# Adherence to methotrexate and associated factors considering social desirability in patients with rheumatoid arthritis: a multicenter cross-sectional study

**DOI:** 10.1101/2022.08.24.22279163

**Authors:** Nobuyuki Yajima, Takashi Kawaguchi, Ryo Takahashi, Hiroki Nishiwaki, Yoichi Toyoshima, Koei Oh, Tsuyoshi Odai, Takayuki Kanai, Donald E. Morisky, Takuhiro Yamaguchi, Tsuyoshi Kasama

**Affiliations:** Division of Rheumatology, Department of Medicine, Showa University School of Medicine, Tokyo, Japan; Department of Healthcare Epidemiology, Kyoto University Graduate School of Medicine and Public Health, Kyoto, Japan; Center for Innovative Research for Communities and Clinical Excellence, Fukushima Medical University, Fukushima, Japan; Department of Practical Pharmacy, School of Pharmacy, Tokyo University of Pharmacy and Life Sciences, Tokyo, Japan; Division of Nephrology, Department of Internal Medicine, Showa University Fujigaoka Hospital, Yokohama, Japan; Showa University Research Administration Center, Tokyo, Japan; Department of Orthopedic Surgery, Showa University Toyosu Hospital, Tokyo, Japan; Department of Orthopedic Surgery, Showa University Northern Yokohama Hospital, Yokohama, Japan; Department of Internal Medicine, Showa University Northern Yokohama Hospital, Yokohama, Japan; Department of Rheumatology, Yokohama Asahi Central General Hospital, Kanagawa, Japan; Department of Nephrology, Kanto Rosai Hospital, Kawasaki, Japan; Department of Community Health Sciences, UCLA Fielding School of Public Health, Los Angeles, United States; Division of Biostatistics, Tohoku University Graduate School of Medicine, Miyagi, Japan

**Keywords:** rheumatoid arthritis, methotrexate, drug monitoring

## Abstract

**Background:** Assessing medication adherence in rheumatoid arthritis (RA) is clinically significant as low adherence is associated with high disease activity. Self-reported medication adherence surveys have been shown to have problems with overestimation of adherence due to social desirability bias. However, no MTX adherence studies adjusted for social desirability have been conducted to date. This study aimed to evaluate adherence to MTX and perform an investigatory search for factors associated with MTX adherence including social desirability.

**Methods:** This cross-sectional multicenter study was conducted among adult RA patients consuming oral MTX for ≥3 months. We examined the distribution of MTX adherence, according to the eight-item Morisky Medication Adherence Scale (MMAS-8). Social desirability was using the Social Desirability Scale (SDS). Furthermore, an exploratory factor analysis involving social desirability was examined to identify factors associated with MTX adherence using linear regression analysis. To deal with missing values, we used multiple imputations with chained equations methods.

**Results:** A total of 165 RA patients were enrolled. The median age was 64 years, and 86.1% were women. Based on the MMAS-8, low, medium, and high adherences were noted in 12.1%, 60.0%, and 27.9% of participants, respectively. High social desirability (coefficient, 0.14; 95% confidence interval [CI], 0.05–0.23; p<0.05) and high age (coefficient per 10 years, 0.16; 95% CI, 0.01–0.03; p<0.05) were associated with high MTX adherence, whereas full-time work was negatively associated with high MTX adherence (coefficient, -0.50; 95% CI, -0.95–-0.05; p<0.05).

**Conclusions:** A large proportion of patients with RA do not take MTX as prescribed. High social desirability, high educational level, and non-full-time work may be associated with high MTX adherence. Physicians should confirm MTX adherence before switching or adding disease-modifying anti-rheumatic drugs in cases of uncontrolled disease activity.

## Background

Rheumatoid arthritis (RA) is a systemic autoimmune disease characterized by chronic inflammation. RA affects the joints and leads to functional disability in the absence of appropriate treatment [1]. The 2015 American College of Rheumatology (ACR) and the 2019 European League against Rheumatism recommendations suggested the early use of disease-modifying antirheumatic-drugs (DMARDs), usually methotrexate (MTX), in most active RA patients [2,3]. Several studies have reported that low medication adherence is associated with high RA disease activity [4,5]. Hence, MTX adherence in RA patients is crucial.

Non-adherence can be categorized as non-initiation, poor execution (accidental or intentional) or non-persistence [6]. As a chronic disease, RA is important for drug continuation, and attention should be focused on non-persistence non-adherence. Several studies on adherence to MTX have been conducted. However, MTX adherence differed among the studies because of the various definitions of adherence, different follow-up durations, and heterogeneity of patient populations. A previous study reported that MTX adherence is associated with age, sex, race, RA disease activity, patient beliefs about the medication, disease duration, mental health, and socio-economic status (SES) [4,7-11]. In a systematic review, beliefs on the necessity and efficacy of MTX, high mood, mild disease, and MTX monotherapy were identified as the most reliable variables related to MTX adherence [12].

The common propensity for people to show themselves in the most favorable light in relation to society values and standards is known as social desirability [13]. Clinically, social desirability may result in underreporting of socially and culturally “undesirable” habits and behaviors, which may eventually result in lost chances for intervention and unrecognized pharmacological contraindications [14,15]. It is well known to influence the use of self-reported questionnaires in research. A previous study reported that social desirability is the tendency to respond to self-reported items in a way that reflects better on the respondent rather than acting accurately and truthfully [16]. Self-reported medication adherence surveys have been shown to have problems with overestimation of adherence due to social desirability bias [17,18]. A previous MTX adherence study has reported the need to control this bias [19]; however, no studies have investigated the MTX adherence survey by controlling for social desirability. Thus, this study aimed to evaluate the effect of already known variables plus social desirability.

## Methods

### Study design

This cross-sectional study was conducted in four hospitals: Showa University Hospital, Showa University Northern Yokohama Hospital, Showa University Fujigaoka Hospital, and Kanto Rosai Hospital. Data collection was conducted only once during the survey.

### Patient population

Participants were outpatients who met the 2010 ACR criteria for RA [20]. The inclusion criteria were age ≥20 years and consumption of oral MTX for ≥3 months. The exclusion criteria were patients with dementia, restlessness, and severe psychiatric disorders. Consecutive sampling was employed. Patients were recruited between August 2013 and October 2014.

### Outcome

The primary outcome was the distribution of MTX adherence according to MMAS-8 at the time of the survey based on the patient’s description. MMAS-8 consists of an eight-item questionnaire [21-23]. MMAS-8 scores range from 0 to 8 and are classified as follows: low, <6; medium, >6 and <8; and high, 8. Originally, MMAS-8 was developed for daily administration of oral medicine. MTX was administered once a week; thus, we developed the Japanese questionnaire for MTX accordingly after obtaining the original developer’s permission. First, a physician (N.Y.), a pharmacist (T.K.) with experience in scale development translated the scale into Japanese. Second, it was back-translated into English by two professional translators. N.Y., T.K. and two professional translators compared the items with the original items, and discussed the questionnaire and achieved a consensus, and revised the translated and back-translated versions. Finally, we were sent to the original author to confirm the semantic content, and some minor improvements were made. The final version was approved by the original author. Then, five Japanese RA participants took part in a pilot test to see if they could understand the questions and respond to them clearly, if the language was clear, if there were any technical or strange words that the subjects couldn’t understand, and if the questions were appropriate for Japanese culture.

### Data collection

Social desirability is typically assessed using the Social Desirability Scale (SDS) developed by Crowne and Marlowe in 1960, with 33 items [24]. By “the urge to seek acceptance by acting in a culturally relevant and acceptable way,” this scale defines social desirability. Furthermore, a 13-item SDS was developed with confirmed validity and reliability [25-27]. In this study, we used the 13-item SDS. The responses are recorded on “True” or “False. Add 1 point to the score for each “True” response to statements 5, 7, 9, 10, and 13. Add 0 points to the score for each “False” response to these statements. Add 1 point to the score for each “False” response to statements 1,2,3,4,6, 8, 11, and 12. Add 0 points to the score for each “True” response to these statements. The score by domains ranges from 0 to 13. Higher scores indicated higher social desirability.

MTX dose, MTX dosing frequency, duration of MTX treatment were collected. RA disease activity was assessed using the Disease Activity Score in 28 joints (DAS28-ESR). [28]. Activities of daily living (ADL) was assessed using the modified Health Assessment Questionnaire (mHAQ), which is a self-reported questionnaire that measures function, including ADL performance. Depression state was defined using the Centre for Epidemiological Studies-Depression (CES-D) scale [29]. The patient’s perceptions towards medications were assessed using the Belief about Medicines Questionnaire-Specific (BMQ-specific), which includes two domains, BMQ-Specific necessity (5 items) and BMQ-Specific concern (5 items). We used the BMQ necessity-concern differential (“BMQ-Specific necessity” minus “BMQ-Specific concern”) predicted adherence most strongly in studies affecting adherence [30]. Pain severity was assessed using the Brief Pain Inventory (BPI). We used the average numerical rating scale (NRS) pain score other clinical studies have applied in the multivariable analysis. Short Form-8 (SF-8) is a health-related quality of life (QoL) questionnaire [31,32]. We used two summary scores: mental component summary (MCS; showing mental status) and physical component summary (PCS; showing physical status). The Japanese versions of mHAQ, CES-D, BMQ, BPI, SDS, and SF-8 were validated [33, 34].

Additionally, we obtained data on SES, including marital status, educational level, employment status, and living status. Data on patient characteristics, medication, and SES were further collected using questionnaires. Other data were obtained from medical chart records. After consulting the doctor, the participants answered the Morisky Medication Adherence Scale (MMAS-8), mHAQ, CES-D, BMQ, BPI, SDS, SF-8, and SES questionnaires; they returned the completed questionnaires to the data center by mail.

### Statistical analyses

MMAS-8, sex, CES-D, and SES were used as categorical variables, whereas age, MTX dose, MTX dosing frequency, duration of MTX treatment, disease duration, DAS28-ESR, mHAQ, BMQ, BPI, SDS, and SF-8 were used as continuous variables. Summary statistics were presented as median with interquartile range (IQR) and numbers with proportion (%). First, we evaluated the distribution of MTX adherence according to MMAS-8. Subsequently, we compared MMAS-8 and pre-described factors using one-way analysis of variance (ANOVA), Kruskal-Wallis test, and chi-squared test. For significant factors, we conducted a trend analysis. Finally, multiple linear regression analysis was performed to exploratively assess factors associated with MTX adherence The co-variables selected were as follows: age, sex, disease duration, RA disease activity (DAS28-ESR), depression state (CES-D), reliability of the medication (BMQ necessity-concern differential), social desirability (SDS), educational level (more than, equal to college-level, or not), and employment status (full-time work or not). These factors were further selected based on previous studies and clinical importance [4,7–11,35-37]. To compare the mean DAS-28 scores among the three groups using ANOVA, the effect size (small, 0.1; medium, 0.25; and large: 0.4) customarily proposed by Cohen [38] was set at medium (0.25), the significance level was set at 5% on both sides, and the power was set at 80%. The total number of cases was calculated to be 159. The target number of cases was 176, assuming a dropout rate of 10%.

To deal with missing values, we used multivariable multiple imputations with chained equations methods to increase power and minimize selection bias since we considered missing data to be an assumption of missing at random. We included age, sex, disease duration, RA disease activity, depression state, reliability of the medication, social desirability, educational level, and employment status for each imputation model. We generated 10 imputed datasets and combined coefficient estimates using Rubin’s rules for each imputation. A two-sided p-value <0.05 was considered statistically significant. All statistical analyses were conducted using STATA 14.2 (StataCorp LP, College Station, TX) software.

### Ethical considerations

The ethics committee of Showa University Hospital (approval number 1446) and Showa University Toyosu Hospital, Showa University Northern Yokohama Hospital, Kanto Rosai Hospital approved this study, and informed consent was obtained from all participants before study enrolment. All study procedures were performed in accordance with the Declaration of Helsinki. Patient information was anonymized and de-identified before analysis.

## Results

### Patient flow chart

Figure 1 shows the patient flow chart. Initially, 181 RA patients were invited. Of 169 patients eligible for the study, four were excluded because of missing MTX adherence data (n=2) and clinical data (n=2). A total of 165 RA patients were included in the final analysis.

**Figure 1.**
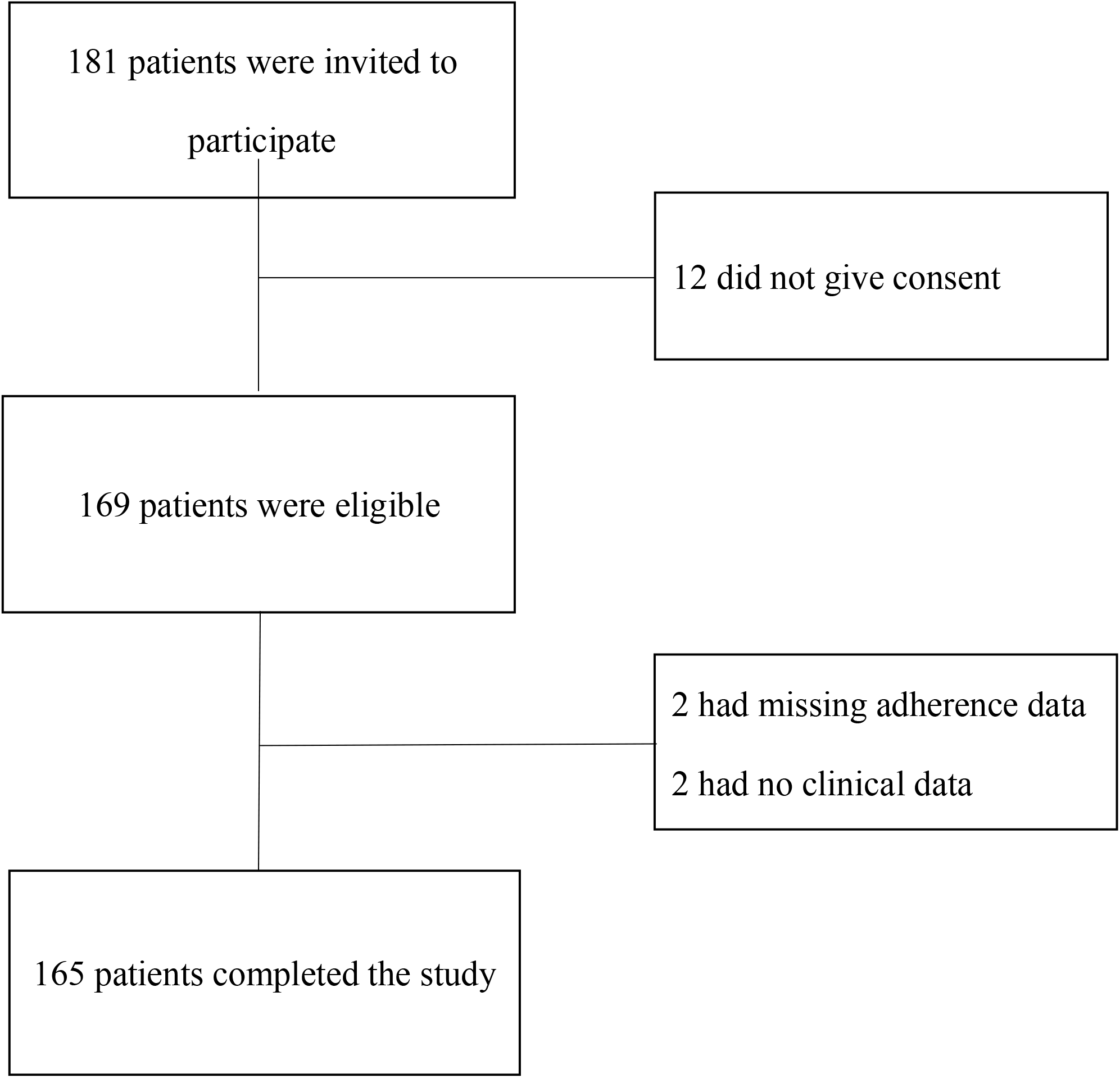
Patient flow chart.

### Baseline characteristics

The characteristics of the participants are shown in Table 1. The median age of the patients was 64 years (IQR, 54–72), and 86.1% were women. The median dose of MTX was 8 mg per week (IQR, 6–12), median MTX dosing frequency was three times per week (IQR, 2–3), and median duration of MTX treatment was 36 months (IQR, 17–75). The proportion of missing data varied from 0 to 15.2% (Supplementary table 1).

**Table 1.**
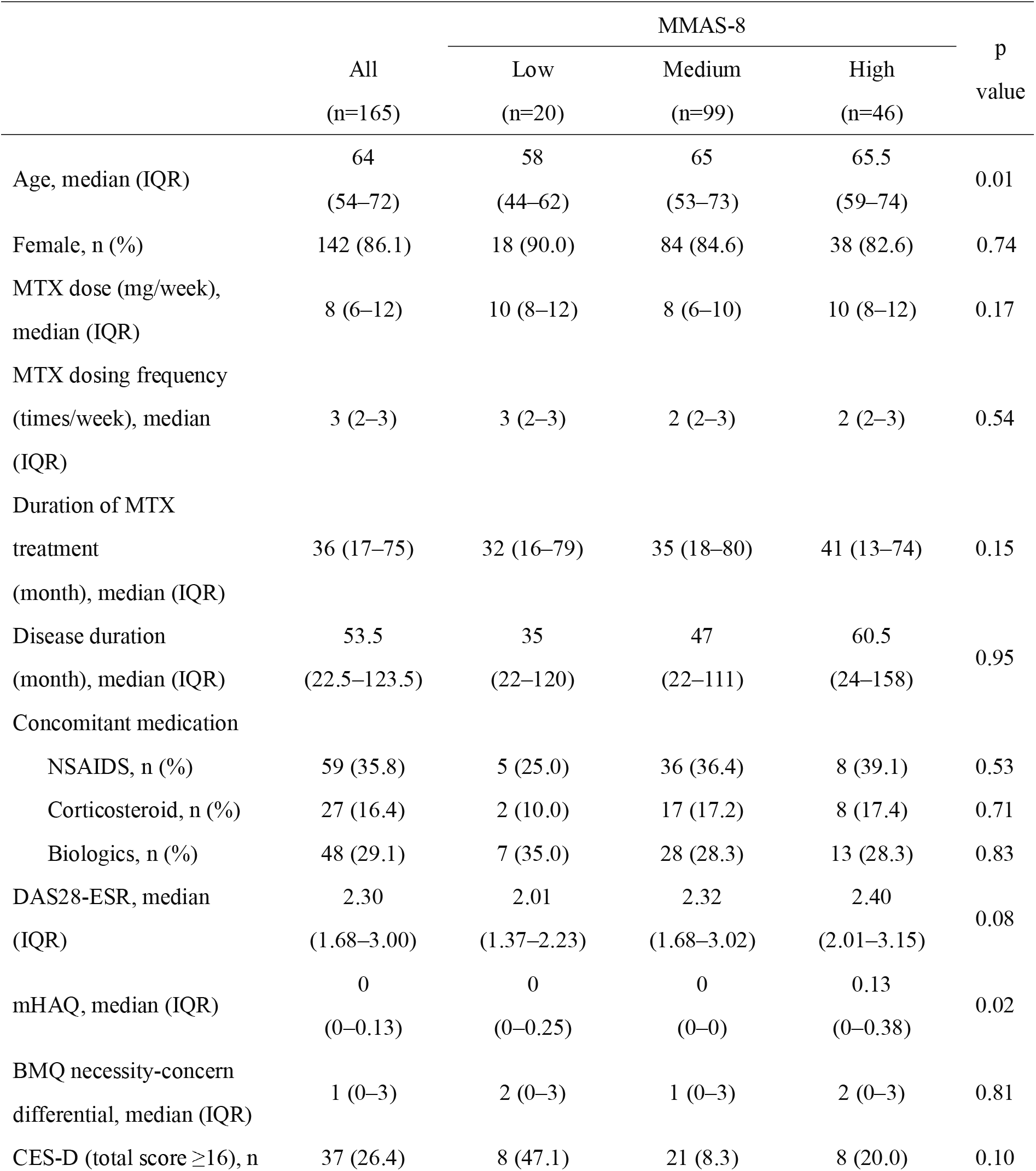
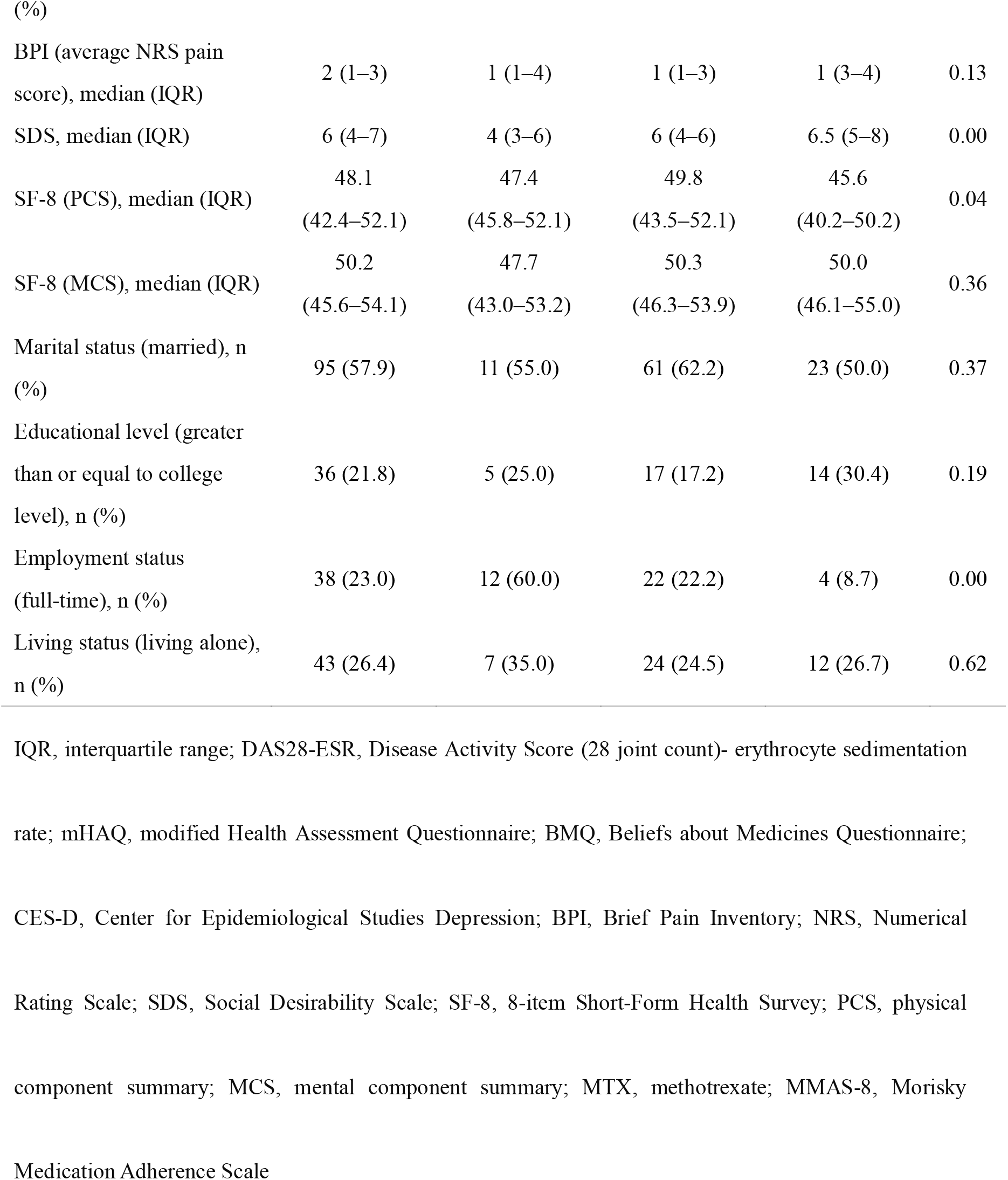
Patient characteristics by grade of MTX adherence divided by MMAS-8 (n=165)

### Distribution of MTX adherence

Based on MMAS-8, low, medium, and high adherence rates were noted in 12% (20/165), 60% (99/165), and 28% (46/165) of cases, respectively. The median MMAS-8 score was 7 (IQR, 6.5–8).

### Patient’s characteristics by grade of MTX adherence

Patient’s characteristics by grade of MTX adherence divided by MMAS-8 are shown in Table 1. A significant correlation between levels of MTX adherence and age, mHAQ, SDS, SF-8 (PCS), and employment status was found. Based on trend analysis, older participants had significantly high adherence, and the higher the social desirability, the higher the adherence. However, a significant correlation between MTX adherence and mHAQ and SF-8 (PCS) was not observed in the trend analysis.

### Factors associated with high MTX adherence considering social desirability

In the multiple linear regression analysis adjusted for age, sex, disease duration, RA disease activity (DAS28-ESR), depression state (CES-D), reliability of the medication (BMQ necessity-concern differential), social desirability (SDS), educational level (more than or equal to college-level or not), and employment status (full-time work or not), higher social desirability (coefficient, 0.14; 95% confidence interval [CI], 0.05–0.23; p<0.05) and higher age (coefficient per 10 years, 0.16; 95% CI, 0.01–0.03; p<0.05) were associated with high MTX adherence, whereas full-time work was negatively related to high MTX adherence (coefficient, -0.50; 95% CI, -0.95–-0.05; p<0.05) (Table 2).

**Table 2.**
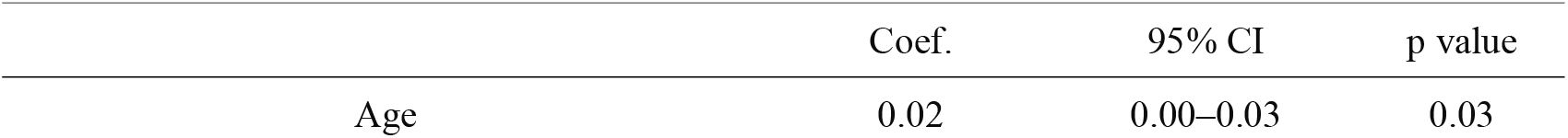
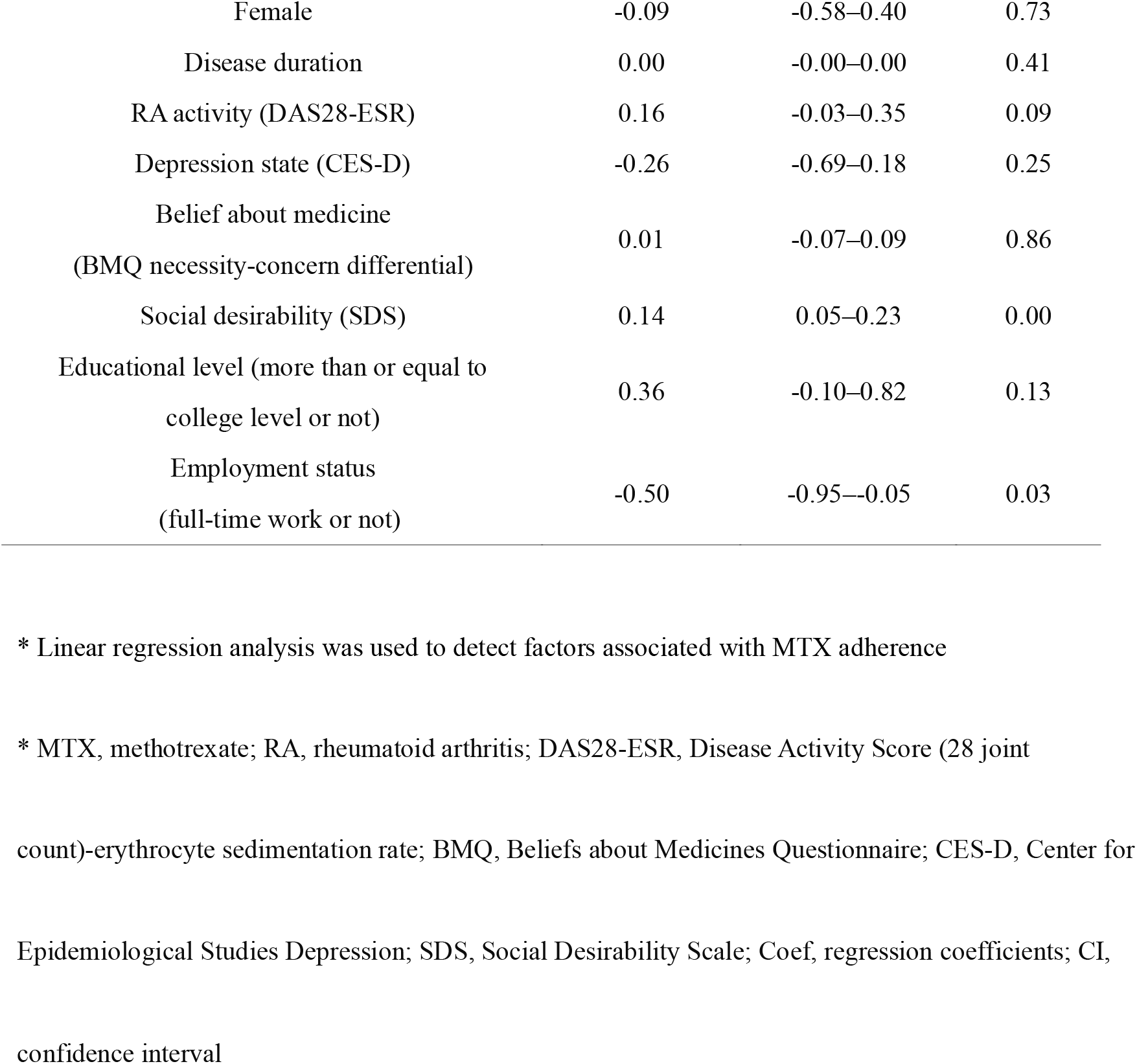
Factors associated with MTX adherence using linear regression analysis (n=165)

## Discussion

Elderly participants and high social desirability were significantly associated with high adherence based on trend analysis. Multiple linear regression analysis showed that high MTX adherence in RA patients is associated with high social desirability, high educational level, and non-full-time work.

In our study, only 27.9% of the participants showed high medication adherence. Several studies reported that adherence to DMARDS was 40%–107% [10,39,40]. There are several potential reasons for the observed low adherence. First, the method of adherence measurement in each study was different. There were interviews, questionnaires, tablet count, drug concentration, and doctor’s judgment as measurements. A previous study showed that the adherence to interview, tablet count, drug concentration, and doctor’s judgment were 96%, 77%, 58%, and 42%, respectively, which showed vast differences [41]. Since each measurement has advantages and disadvantages, the gold standard is not determined. We chose a self-reported questionnaire in this study since this scale is not expensive or invasive and can be possibly validated. Second, the time point of adherence measurement was different between the studies. We measured adherence during the maintenance period (mean disease duration, 53 months). A previous report suggested that the adherence of DMARDs in the early period was low (58%) [5]. Another study reported the adherence to salazosulfapyridine as 87% at three years from the start of medication [36]. In contrast, the adherence to DMARDs in the maintenance period varied from 84%,^4^ 70% [42], to 95.4% [43]. Previous reports have suggested that the adherence to DMARDs was not different between the early and maintenance periods. It is crucial to note that the time point of measuring medication adherence is the maintenance period because RA is a chronic disease that requires DMARDs for a long duration. The third is the difference in participant characteristics. Living status and marital status affected adherence [11, 44]. Our study could not adjust the living status and marital status because we could not include more factors based on our sample size. In addition, the age of the participants may have had an effect. In the original MMAS-8 article [21], the mean age was 52 years; however, in our study, the median age was 64 years, which means that the participants in our study were older, and this may have influenced the results. Furthermore, older people may not have revealed the truth. Thus, when referring to drug adherence studies, the method of measuring adherence, whether the target patient is in the disease’s early or maintenance phase, and the patient’s characteristics should be confirmed before adaptation.

Our study detected high social desirability, high educational level, and non-full-time work as factors affecting MTX adherence. A previous study reporting MTX adherence found that adherence is associated with age, sex, race, RA disease activity, patient beliefs about the medication, disease duration, mental health, and SES [4,7-11,35-37]. The discrepancy between our findings and those of other studies could be attributed to three reasons. First, we measured and adjusted social desirability, which possibly affected the answers in the questionnaire. Although a recent MTX adherence research mentioned the necessity to account for this bias [19], no studies have examined how MTX adherence survey accounts for social desirability. The results may have differed since we conducted a factor search that included social desirability. Second, racial differences may have led to different outcomes. Our study was based on a Japanese population; however, previous studies were mainly on white people, black people, and Hispanic people. Canon et al. reported that the adherence to MTX was associated with the Caucasian race [4]. Therefore, adherence may be influenced by race. Third, the definition of medication adherence varies between studies. We used MMAS-8 in our study. Conversely, previous studies used other methods to assess medication adherence, such as physicians’ estimation, different self-reported scales, drug concentration, tablet count electronic monitors, and other questionnaires. A prior study reported that medication adherence differs because the measurement method employed seems to vary [12]. Therefore, we need to practice caution when using research results because the results may differ depending on the method of measuring adherence and the variables incorporated, including social desirability.

Our study had three strengths. First, we assessed social desirability, leading to social desirability bias. To our knowledge, no clinical studies adjusting social desirability have investigated the association between MTX adherence and various factors. Second, to our knowledge, this is the first research to examine MTX adherence in an Asian population. Third, the response rate to the questionnaires was high (93.4%), thereby strengthening internal validity.

This study has significant implications for the clinician. Our findings may prevent unnecessary DMARDs changes since physicians tend to overestimate patients’ medication adherence [45]. They should confirm MTX adherence before switching or adding DMARDS. Accordingly, this provides an opportunity to reduce the healthcare cost and adverse events of medication.

However, this study had several limitations. First, in the multivariable analysis, some immeasurable essential factors (such as characteristics of each participant, joint deformities, number of medications, and MTX dosing frequency) that could affect MTX adherence possibly exist. We did not determine patient personality due to the difficulty in quantifying it. In addition, we did not collect radiographs even if they are necessary for determining joint deformity because they are burdensome for patients. However, we adjusted for joint deformity using the mHAQ to reduce its possible influence on the findings. In addition, although the number of MTX doses was collected, it was not possible to incorporate variables due to sample size limitations. Second, the generalizability of our findings is limited. Our study setting was primarily university hospitals in Japan; thus, the results of this study cannot be applied to the clinical setting, in small- and medium-sized hospitals, or in settings including other races. Furthermore, the highest dose of MTX in Japan in the study was 8 mg per week. This dose is less than the standard dose of 20 mg worldwide, which limits the indications. Third, we used a self-reported questionnaire of adherence. Hence, the completed questionnaire possibly influenced social desirability. Although we could not adjust this bias completely, social desirability was added as a factor for analysis. Furthermore, MMAS-8 is “self-perceived adherence” and not true drug adherence. Therefore, even a factorial search including social desirability may not reveal the truth. Forth, The Marlowe-Crowne Social Desirability scale may have inadequate validity and reliability [46]. The validity of this scale has not been examined in the target population of RA patients. There are also negative studies on the reliability of the SDS scale. As a future study, we would like to conduct a validity study of this scale in RA patients.

## Conclusions

Our results demonstrated that sixty percent of the RA patients had moderate adherence to MTX. Moreover, high MTX adherence in RA patients is associated with high social desirability, high educational level, and non-full-time work.

## Data Availability

The dataset analyzed in this paper is available from the corresponding author on reasonable request.

## List of Abbreviations

ADL: Activities of Daily Living
BMQ: Beliefs about Medicines Questionnaire
BPI: Brief Pain Inventory
CES-D: Center for Epidemiological Studies Depression
DAS28-ESR: Disease Activity Score (28 joint count)-erythrocyte sedimentation rate
DMARDs: disease-modifying antirheumatic-drugs
MCS: mental component summary
mHAQ: modified Health Assessment Questionnaire
MMAS-8: Morisky Medication Adherence Scale
MTX: methotrexate
NRS: Numerical Rating Scale
PCS: physical component summary
RA: rheumatoid arthritis
SDS: Social Desirability Scale
SF-8: 8-item Short-Form Health Survey

## Declarations

### Ethics approval and consent to participate

The ethics committee of Showa University Hospital (approval number 1446) and other each institution approved this study, and informed consent was obtained from all participants before study enrolment. All study procedures were performed in accordance with the Declaration of Helsinki. Patient information was anonymized and de-identified before analysis.

### Consent for publication

Not applicable.

### Availability of data and materials

About MMAS8

The use of the MMAS diagnostic adherence assessment instrument is protected by US copyrighted and trademarked laws. Permission for use is required. A license is available from MORISKY MEDICATION ADHERENCE RESEARCH, LLC., Donald E. Morisky, ScD, ScM, MSPH, MMAR, LLC, 294 Lindura Ct., Las Vegas, NV 89138.

### Authors’ contributions

Nobuyuki Yajima: Methodology, Project administration, Writing – original draft, Writing – Review & Editing

Takashi Kawaguchi: Supervision, Conceptualization, Project Administration, Writing – original draft, Writing – Review & Editing

Ryo Takahashi: Data curation, Writing – original draft, Writing – Review & Editing

Hiroki Nishiwaki: Data curation, Writing – original draft, Writing – Review & Editing

Youichi Toyoshima: Data curation, Writing – original draft, Writing – Review & Editing

Koei O: Data curation, Writing – original draft, Writing – Review & Editing

Tsuyoshi Odai: Data curation, Writing – original draft, Writing – Review & Editing

Takayuki Kanai: Data curation, Writing – original draft, Writing – Review & Editing

Donald E. Morisky: Supervision, Writing – original draft, Writing – Review & Editing

Takuhiro Yamaguchi: Formal Analysis, Supervision, Writing – original draft, Writing – Review & Editing

Tsuyoshi Kasama: Supervision, Writing – original draft, Writing – Review & Editing

## Acknowledgments

We thank Noboru Murata (Kikuna Memorial Hospital) and Osamu Namiki for critically reviewing the manuscript.

