## Supplementary material for "Adherence to methotrexate and associated factors considering social desirability in patients with rheumatoid arthritis: a multicenter cross-sectional study": table1._Missing_data_of_patient_s_characteristics (from another version of this preprint: version 1, 2022-08-25)

**Supplementary table 1. Missing data of patient characteristics**

|  | Missing data, n | | |
| --- | --- | --- | --- |
|  | MMAS-8  Low  (n=20) | MMAS-8  Medium  (n=99) | MMAS-8  High  (n=46) |
| Age | 0 | 0 | 0 |
| Female | 0 | 0 | 0 |
| MTX dose | 0 | 0 | 0 |
| MTX dosing frequency | 0 | 0 | 0 |
| Duration of MTX treatment | 0 | 0 | 1 |
| Disease duration | 1 | 0 | 0 |
| Concomitant medication | 0 | 0 | 0 |
| NSAIDs, n (%) | 0 | 0 | 0 |
| Corticosteroid, n (%) | 0 | 0 | 0 |
| Biologics, n (%) | 0 | 0 | 0 |
| DAS28-ESR | 2 | 7 | 3 |
| mHAQ | 1 | 2 | 1 |
| BMQ (necessity-concern) | 1 | 3 | 3 |
| CES-D | 3 | 16 | 6 |
| BPI (average NRS pain score) | 1 | 1 | 2 |
| SDS | 1 | 8 | 6 |
| SF-8 (PCS), median (IQR) | 0 | 2 | 0 |
| SF-8 (MCS), median (IQR) | 0 | 2 | 0 |
| Marital status | 0 | 1 | 0 |
| Educational level | 0 | 0 | 0 |
| Employment status | 0 | 0 | 0 |
| Living status | 0 | 1 | 1 |

MMAS-8, Morisky Medication Adherence Scale; MTX, methotrexate; DAS28-ESR, Disease Activity Score (28 joint count)-erythrocyte sedimentation rate; mHAQ, modified Health Assessment Questionnaire; BMQ, Beliefs about Medicines Questionnaire; CES-D, Center for Epidemiological Studies Depression; BPI, Brief Pain Inventory; SDS, Social Desirability Scale; SF-8, 8-item Short-Form Health Survey; PCS, physical component summary; MCS, mental component summary; IQR, interquartile range; NSAIDs, non-steroidal anti-inflammatory drugs
